# Characteristic of IgA and IgG antibody response to SARS-CoV-2 infection in an Italian referral Covid-19 Hospital

**DOI:** 10.1101/2020.11.16.20232470

**Authors:** Carnicelli Annamaria, Fiori Barbara, Ricci Rosalba, Piano Alfonso, Bonadia Nicola, Taddei Eleonora, Fantoni Massimo, Murri Rita, Cingolani Antonella, Barillaro Christian, Cutuli Salvatore Lucio, Marchesini Debora, Della Polla Davide Antonio, Forte Evelina, Fuorlo Mariella, Di Maurizio Luca, Amorini Paola, Franceschi Francesco, Sanguinetti Maurizio

## Abstract

**Introduction:** Antibody response play a fundamental role in the natural history of infectious disease. A better understanding of the immune response in patients with SARS-CoV-2 infection could be important for identifying patients at greater risk of developing a more severe form of disease and with a worse prognosis.

**Methods:** We performed a cross-sectional analysis to determine the presence and the levels of both anti-SARS-CoV-2 IgG and IgA in a cohort of hospitalized patients with confirmed infection at different times in the natural history of the disease. Patients enrolled when admitted at the emergency department were prospectively followed up during hospital stay.

**Results:** Overall, 131 patients were considered with a total of 237 samples processed. Cross-sectional analysis showed that seroconversion for IgA seems to occur between days 5 and 15 while IgG response seems to occur slightly later, peaking at day 20 after symptoms onset. Both IgA and IgG were maintained beyond two months. Severe patients showed a higher IgA response compared with mild patients when analyzing optical density (8.3 versus 5.6, p < 0.001). Prospective analysis conducted on 55 patients confirmed that IgA appear slightly earlier than IgG. After stratifying for the severity of disease, both the IgA and IgG response was more vigorous in severe cases. Moreover, while IgG tended to stabilize, there was a relevant decline after the first month of IgA levels in mild cases.

**Conclusion:** IgA and IgG antibody response is closely related although seroconversion for IgA occurs earlier. Both IgA and IgG are maintained beyond two months. Severe patients showed a more vigorous IgA and IgG response. IgA levels seem to decline after one month since the onset of symptoms in mild cases.

## Introduction

At the end of December 2019, a cluster of patients suffering from a new kind of pneumonia of unknown etiology has been observed in Wuhan (China) [1]; subsequently, it was attributed to a new virus (SARS-CoV-2), a beta-coronavirus phylogenetically similar to the causative agent of SARS, which determines the respiratory disease then called COVID-19[2].

From December 2019 to today, the spread of SARS-CoV-2 infection has taken on the size of a pandemic[3]. According to the latest WHO data, to date, more than 38 million cases and one million deaths from COVID-19 have been confirmed worldwide [4], challenging the healthcare systems all over the world.

Moreover, although mortality from SARS-CoV-2 was found to be lower than that from SARS and MERS, COVID-19 deaths are greater in absolute number due to the higher transmissibility of this new virus[5].

As a result, public health services around the world have implemented different strategies for rapid identification of both infected and asymptomatic carriers in order to prevent virus transmission.

To date, direct detection of SARS-CoV-2 virus RNA throughout real-time reverse-transcriptase–polymerase-chain-reaction (rtPCR) assay from specimen of upper or lower airways has become the standard method for diagnosing SARS-CoV-2 infection in microbiology laboratories worldwide, although these tests have several limitations and many false negatives have been reported[6].

Recently, detection of antibodies specific to SARS-CoV-2 has also been recognized as deterministic evidence for confirmed SARS-CoV-2 infection[6]. However, the role and the pattern of antibody response in SARS-CoV-2 infection is not yet fully understood and many gaps are still present[7].

Neutralizing antibodies play a fundamental role in the clearance of viruses and are used as a gold standard for the evaluation of immunization in many viral infections.

Previous studies carried out during the SARS epidemic showed that the antibody titer and the rate of seroconversion were associated with the severity of the disease. In particular, in the study conducted by Lee et al.[8], seroconversion was observed around the sixteenth day and the peak of IgG was reached at 4 weeks; moreover, the rapidity of seroconversion (<16 days) was associated with the need for hospitalization in intensive care unit (ICU), while high IgG antibody titers were associated with the need for ventilation and hospitalization in ICU. Other studies carried out during SARS epidemic, showed that patients with milder forms of disease had lower antibody titers[9–12], suggesting that, as observed in other infectious diseases (such as Dengue), an important humoral response may be associated with an exaggerated immune response and generate a cytokine storm[13–16], which can contribute to disease progression.

Despite many similarities to SARS-CoV infection, little information is currently available on the role of the antibody response in the evolution of SARS-CoV-2 disease. Moreover, most of the studies available at the moment explore the characteristic only of IgM and IgG response to SARS-CoV-2 infection[17–20], while little information are available about all the other mechanisms involved in the immune response, such as, for example, the role of IgA antibodies.

In the past, several studies have pointed out the crucial role of IgA antibodies as an effective defense against respiratory infections[21] and previous studies on SARS-CoV have shown similar kinetics of IgA, IgM, and IgG[13,22].

A better understanding of the immune response in patients with SARS-CoV-2 infection could be important for the development of an effective vaccine and for identifying patients at greater risk of developing a more severe form of disease and with a worse prognosis[17].

In our study we investigate the characteristics of IgA and IgG antibody response in a cohort of patients with confirmed infection from SARS-COV-2 admitted to the Emergency Department (ED) of a referral COVID Hospital in Italy.

## Methods

We conducted an observational, single-center study in a tertiary ED of a university hospital located in Rome, Italy, currently serving as a referral center for COVID-19.

### Aim the of the study

Aim of the study was to describe the pattern of IgG and IgA antibody response against SARS-CoV-2 in hospitalized individuals.

The study involved two kind of analysis:

1. A cross-sectional analysis which had the aim to evaluate the presence and levels of specific IgG and IgA antibodies at different times in the natural history of the disease in patients referred to the ED or hospitalized in our institution with confirmed SARS-CoV-2 infection in a considered time frame;
2. A prospective observational analysis with the aim to evaluate among patients referred to the ED with confirmed SARS-CoV-2 infection, the presence and level of IgA and IgG antibodies at the time of diagnosis and at different time points after the onset of symptoms.

Furthermore, we assessed whether antibody levels are associated with the severity of infection.

### Patient enrollment and study population

Enrollment procedures for both parts of the study are detailed below.

For the prospective analysis all patients presenting to our institution’s ED between May, 1^st^ and May, 31^st^ who had a clinical suspicion of COVID-19 were evaluated for inclusion in the study; patients who fulfilled the inclusion criteria were enrolled in the study; for each patient, the day of symptoms onset, as recalled by the patient, was registered and a blood sample for IgA and IgG measurement was drawn.

Subsequently, patients hospitalized in any of the COVID-19-dedicated wards of our institution (either in COVID-19-dedicated general medical wards, in COVID-19-dedicated ICU or in our institution’s supervised residential care) underwent blood draw during the course of their illness at 7, 14, 21 days, 1 and 2 months since the admission.

Such data were analyzed independently from data obtained for the cross-sectional part of the study. For the cross-sectional analysis, all patients who were hospitalized between May 1^st^ and May 31^st^ in any of the COVID-19-dedicated wards of our institution (either in the ED, in COVID-19-dedicated general medical wards, in COVID-19-dedicated ICU or in our institution’s supervised residential care) were evaluated for inclusion in the present study; patients fulfilling enrollment criteria underwent a single blood draw for IgA and IgG testing. For each patient, the date of symptoms onset was recorded.

All data point obtained for the prospective enrollment were pooled with those obtained for the cross-sectional enrollment before data analysis.

Inclusion criteria were the following:

- suggestive clinical presentation (dyspnea, fever, cough, coryza; for patients with a pre-existing chronic respiratory condition, worsening dyspnea or worsening respiratory failure; chest imaging at X-ray, CT or lung ultrasound of interstitial involvement or multiple consolidations suspected for COVID-19);
- microbiological confirmation of the diagnosis of SARS-CoV2 infection, defined as direct detection of SARS-CoV-2 virus RNA throughout rtPCR assay on nasopharyngeal swab performed at the time of access to the ED, in the preceding 7 days or during the first 7 days of the index hospitalization;
- patient was 18 years or older at the time of the ED admission;
- patient was willing to participate in the study.

Exclusion criteria were

- primary or secondary immunodeficiencies;
- active hematological malignancy;
- lack of microbiological confirmation of SARS-CoV2 infection during the index hospitalization;
- patients who were asymptomatic or in whom it was not possible to clearly define the onset of symptoms.

### Study Examinations

Blood specimen were collected via venipuncture in a 9ml dry test tube, after discarding the first 2ml of blood. The variables collected for each patient were the following: age, sex, clinical symptoms, time of onset of symptoms, radiological investigations (Chest X-ray, Chest CT scan), rt-PCR for SARS-CoV-2 on oro-pharyngeal and naso-pharyngeal swab (and bronchoalveolar lavage if performed), laboratory tests (Hb, MCV, WBC, neutrophil and lymphocyte count, C-reactive protein, fibrinogen, d-dimer, LDH), need for ICU admission, outcome of the hospital stay (death or discharge), overall severity of SARS-CoV-2 infection (asymptomatic, mild disease, interstitial pneumonia without respiratory failure, mild-moderate-severe ARDS). ARDS and ARDS severity was defined according to generally used international guidelines[23,24]. This study was conducted in accordance with the principles of Helsinki declaration[25].

### Assay of IgA and IgG antibodies

Serum was separated by centrifugation at 2500g for 5 min within 12 h of collection. The specimens were analyzed with SARS-CoV-2 IgG and IgA kits (Euroimmun, Lübeck, Germany;www.euroimmun.com) on the lab workstation platform. The EUROIMMUN Anti-SARS-CoV-2 Assay is an enzyme-linked immunosorbent assay (ELISA) that provides semi-quantitative in vitro determination of human antibodies of immunoglobulin classes IgA and IgG against SARS-CoV-2 in serum or EDTA plasma. Each kit contains microplate strips with 8 break-off reagent wells coated with S1 domain of viral spike protein recombinant of SARS-CoV-2. In the first reaction step, diluted patient samples are incubated in the wells. In the case of positive samples, specific antibodies will bind to the antigens. To detect the bound antibodies, a second incubation is carried out using an enzyme-labelled antihuman IgA or IgG (enzyme conjugate) catalyzing a color reaction. Results are evaluated semi-quantitatively by calculation of a ratio of the extinction of the control or patient sample over the extinction of the calibrator. This ratio is interpreted as follows: < 0.8 negative; ≥ 0.8 to < 1.0 borderline; ≥ 1.1 positive.

### Statistical analysis

We planned to perform an exploratory data analysis on levels of circulating antibodies at different time points and on the proportion of patients with positive serological assays at different time points.

First, we divided the same samples in 5-day time periods and we calculated the proportion of positive patients for each time frame. To detect the most likely time of seroconversion for each antibody class, we compared the proportion of positive patients for each time frame with the proportion of positive patients of the subsequent time frame, adjusting significant p values for multiple comparison with the Holm method.

To assess the possible association between antibodies level or antibodies development and severity of disease, we divided our blood samples according to the maximum severity of the disease. Patients who developed a moderate or severe ARDS were classified as having severe disease, while other patients were classified as having mild disease. We compared the rate of positive sample for each class of antibodies between severe and mild patients both overall and after stratifying by time frame since the onset of symptoms. For this analysis, the positive/negative result of the antibody detection assay was considered as a dichotomous variable and each comparison was performed using the Pearson’s chi-square test or the Fisher exact test, as appropriate. Multiple comparison correction was performed only when a statistically significant result was found using the Holm procedure. Statistical significance was considered as an alpha level of 0,05, two-sided.

The degree of antibody response was also compared between patients with severe or mild disease, using the optical density of the sample as a surrogate for the antibody titer. For this analysis, severe and mild patients were compared both overall and after stratifying for the above-defined time frames. Given that optical density is a continuous variable, either the Student’s t-test or the Wilcoxon-Mann-Whitney test were used, as appropriate. Multiple comparison correction was performed only when statistically significant result was found using the Holm procedure. Statistical significance was considered as an alpha level of 0,05, two-sided.

It should be noted that this is a descriptive study; thus, calculated p-values are only provided to identify most pronounced differences and are not meant to quantify the risk of alpha error on hypothesis testing.

Data collection was performed using Microsoft Excel for Office 365 (Microsoft Corporation, Redmond, WA, USA). Statistical analysis and plot design were performed with the R statistical language version 4.0.2[26] via the RStudio IDE (RStudio, PBC, Boston, MA, USA)[27], using the Tidyverse package[28].

## Results

### Patient population and serological samples

Initially, 218 patients with suspected SARS-COV-2 infection were considered for inclusion. Of those, 30 were excluded because they did not have a microbiological confirmation of SARS-CoV-2 infection; Among 188 patients with confirmed infection, 20 patients were excluded because they were asymptomatic and 37 patients were excluded because the onset of symptoms could not be clearly identified.

Overall, serological samples included in the analysis were obtained from 131 patients (see Table 1). 55 patients had multiple samples drawn during their hospital stay. The median age of our patients was 64 years (IQR 51.5 – 75) (). 34 patients developed a severe form of the disease (26%), while 97 were classified as having a mild form (74%). The median time between symptoms onset and sample collection was 23 days (IQR 12-45, range 1-88).

**Table 1.** Descriptive characteristics of the patients enrolled in the study

Appendix: Tables and figures
| Variable | Measure | IQR or percentage |
| --- | --- | --- |
| Number of patients | 131 |  |
| Age | 64 | 51,5 - 75 |
| Female | 29 | 22% |
| Male | 109 | 78% |
| **Disease severity** |  |  |
| Severe | 34 | 26% |
| Mild | 97 | 74% |
| **Outcome** |  |  |
| Death | 15 | 11% |
| Discharged home | 101 | 77% |
| Still hospitalized | 15 | 11% |

Overall, there were a total of 237 samples processed. Each sample was processed both for IgA and for IgG antibodies. Of those, 183 resulted positive for IgA (77.2%) and 174 for IgG (73.4%).

### Cross-sectional analysis

#### Positivity rate per time frame: IgA

The rate of positive samples for IgA divided per time frame are shown in Table 2.

**Table 2.** Positive rate for IgA and IgG for different time frames since the onset of symptoms in the cross-sectional analysis. P-value are calculated for comparison between each time frame and the subsequent one.

| Time Frame (in days) | Number of patients | IgA |  |  |  |  |  | IgG |  |  |  |  |  |
| --- | --- | --- | --- | --- | --- | --- | --- | --- | --- | --- | --- | --- | --- |
|  |  | Positive number | Positive rate | Lower 95% CI | Upper 95% CI | Unadjusted p | Adjusted p (Holm) | Positive number | Positive rate | Lower 95% CI | Upper 95% CI | Unadjusted p | Adjusted p (Holm) |
| 0-5 | 18 | 3 | 0,167 | 0,044 | 0,423 | 1,000 | 1,000 | 2 | 0,111 | 0,019 | 0,361 | 1,000 | 1,000 |
| 6-10 | 18 | 3 | 0,167 | 0,044 | 0,423 | 0,004 | 0,044 | 2 | 0,111 | 0,019 | 0,361 | 0,229 | 1,000 |
| 11-15 | 17 | 12 | 0,706 | 0,440 | 0,886 | 1,000 | 1,000 | 6 | 0,333 | 0,144 | 0,588 | 0,026 | 0,290 |
| 16-20 | 18 | 12 | 0,667 | 0,412 | 0,856 | 0,500 | 1,000 | 13 | 0,765 | 0,498 | 0,922 | 1,000 | 1,000 |
| 21-25 | 17 | 14 | 0,824 | 0,558 | 0,953 | 0,945 | 1,000 | 13 | 0,722 | 0,464 | 0,893 | 0,447 | 1,000 |
| 26-30 | 18 | 16 | 0,889 | 0,639 | 0,981 | 0,492 | 1,000 | 15 | 0,882 | 0,623 | 0,979 | 0,441 | 1,000 |
| 31-35 | 17 | 17 | 1,000 | 0,771 | 1,000 | 1,000 | 1,000 | 18 | 1,000 | 0,781 | 1,000 | 0,977 | 1,000 |
| 36-40 | 18 | 17 | 0,944 | 0,706 | 0,997 | 0,554 | 1,000 | 16 | 0,941 | 0,692 | 0,997 | 0,638 | 1,000 |
| 41-45 | 17 | 14 | 0,824 | 0,558 | 0,953 | 0,945 | 1,000 | 15 | 0,833 | 0,577 | 0,956 | 1,000 | 1,000 |
| 46-50 | 18 | 16 | 0,889 | 0,639 | 0,981 | 1,000 | 1,000 | 15 | 0,882 | 0,623 | 0,979 | 0,959 | 1,000 |
| 51-55 | 17 | 16 | 0,941 | 0,692 | 0,997 | 0,977 | 1,000 | 17 | 0,944 | 0,706 | 0,997 | 1,000 | 1,000 |
| 56-60 | 18 | 18 | 1,000 | 0,781 | 1,000 | ns | ns | 17 | 1,000 | 0,771 | 1,000 | ns | ns |
| > 60 | 26 | 26 | 1,000 | 0,822 | 1,000 |  |  | 26 | 1,000 | 0,822 | 1,000 |  |  |

There were 18 samples drawn in the first 5 days from the onset of symptoms. Of those, 3 tested positive for IgA (point estimate: 16,7%, 95% CI 4.4 – 42.3%). Similarly, there were 18 samples drawn between day 6 and day 10, and of those only 3 tested positive for IgA (16,7%, 95% CI 4.4 – 42.3%). Conversely, there were 17 samples drawn between day 11 and day 15 and, of those, 12 tested positive for IgA (70.6%, 95% CI 44.0-88.6%). The point estimate for the proportion of positive samples remained stable over 80% from day 21 onward. There were 26 samples drawn beyond 60 days after the onset of symptoms, all of which tested positive for IgA (100%, 95% CI 82.2 – 100%).

The highest increase in rate of proportion of positive samples was observed between the second and the third time frame, that is between the 6-10 days samples and the 11-15 days samples.

Difference between each time frame and the subsequent one showed a peak for samples drawn between days 11 and 15, suggesting that IgA response develop in the majority of patients between day 5 and 15 since the beginning of symptoms (Figure 1).

**Figure 1.**
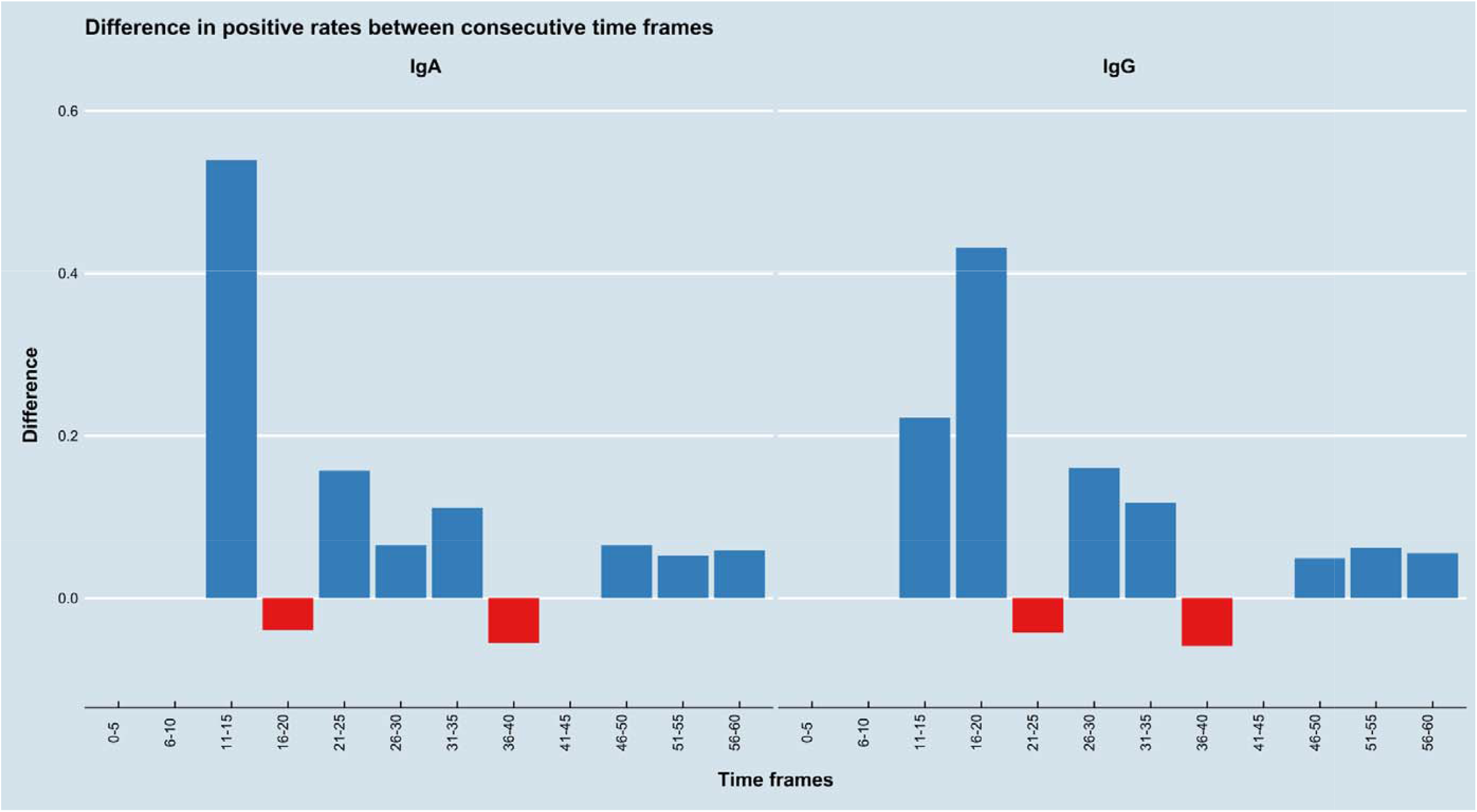
Positivity rates shows a more abrupt increase for IgA at the 11-15 time frame, while there is a more smoldered trend for IgG. Each bar represents the difference in positivity rates between each time frame and the subsequent one.

#### Positive rate per time frame: IgG

The rate of positive samples for IgG divided per time frame are given in Table 2.

There were 18 samples drawn in the first 5 days since the onset of symptoms. Of those, 2 tested positive for IgG (11.1%, 95% CI 1.9 – 36.1%). A similar result applies to the samples drawn between days 6 and 10. A first increase for the rate of positive samples was observed for the samples drawn between 11 and 15 days from the onset of symptoms. At this time frame, there were 18 samples and of those, 6 tested positive for IgG (33.3%, 95% CI 14.4 – 58.8%). A second increase in the rate of positive samples was observed for samples drawn between 16 and 20 days. At this time frame, there were 17 samples and, of these, 13 tested positive for IgG (76.5%, 95% CI 49.8 – 92.2%). Of note, the difference between positive rates at time frame 11-15 and 16-20 was the only difference to reach statistical significance, although this significance was not maintained after correction for multiple comparison with the Holm method. There were 26 samples drawn beyond 60 days from the onset of symptoms, all of which tested positive for IgG (100%, 95% CI 82.2 – 100%).

Differences between consecutive time frames showed a more marked increase in the rate of positive values for samples drawn between days 11 and 15 and for samples drawn between days 16 and 20, suggesting that appearance of IgG may happen between days 6 and 20 (see Figure 1).

#### Optical density

Overall, there were 237 samples tested for IgA. The median optical density was 4.7 unit (IQR 1.5 – 8.3). The same 237 samples were also tested for IgG, for which the median optical density was 5.5 (IQR 0.5 – 9.1). Visual inspection of the dot-plot of optical density versus time showed a similar trend in time for both IgA and IgG (Figure 2).

**Figure 2.**
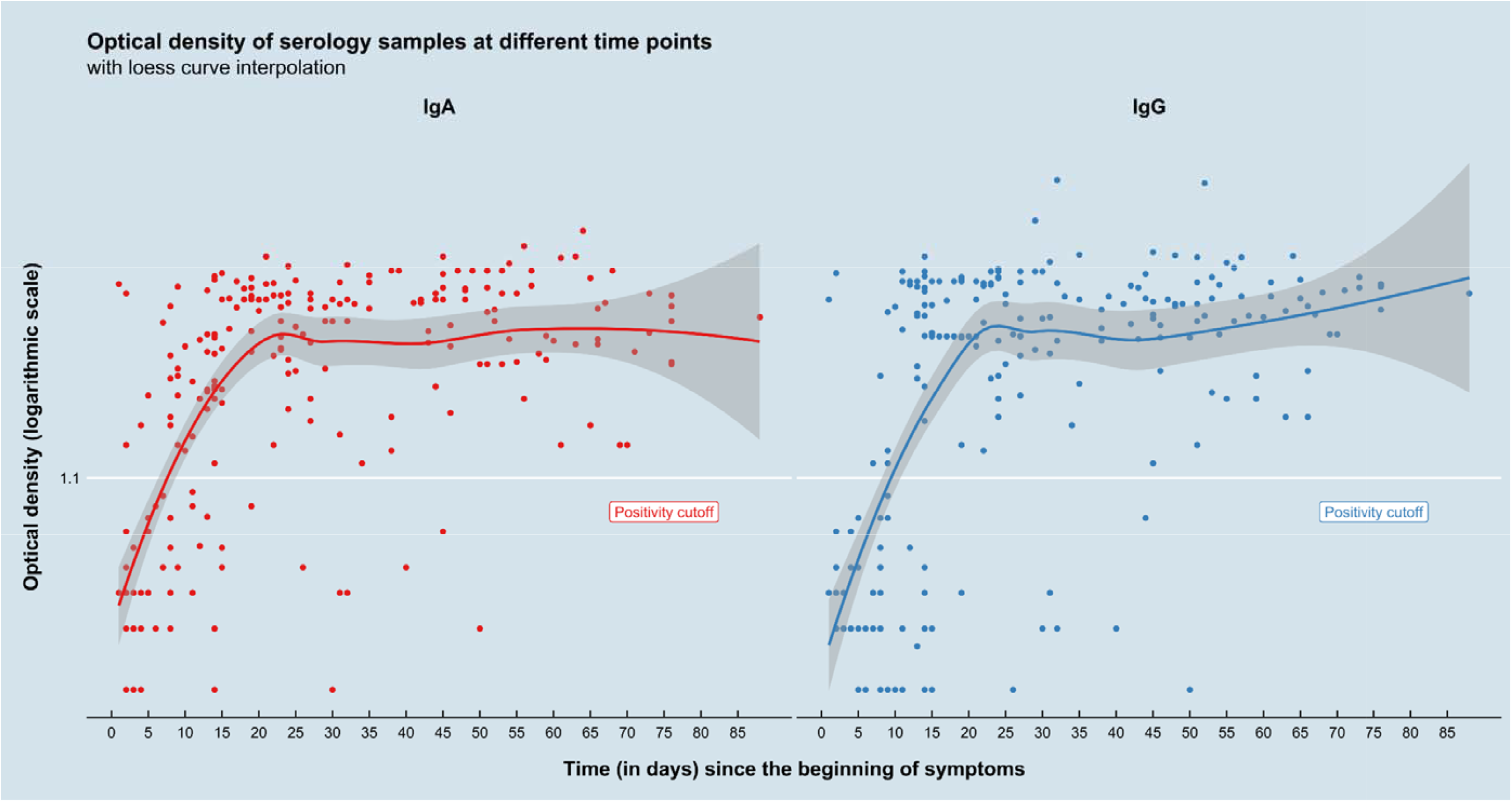
Optical density in time for all samples divided by Ig class

#### Serological response according to severity of disease

After stratifying per time frame and per severity of the disease, there was no statistically significant difference between severe patients and mild patients in the rate of positive result.

Conversely, there was a statistically significant difference for the overall optical density of IgA between severe patients and mild patients, with severe patients overall showing a higher optical density for IgA (8.3 versus 5.6, p < 0.001). A smaller difference was observed for IgG, with more severe patients showing an overall higher optical density, although for IgG statistical significance was not reached (8.4 versus 7.0, p = 0.09) (Figure 3). Such a difference was not observed after stratifying the samples per time since the onset of symptoms.

**Figure 3.**
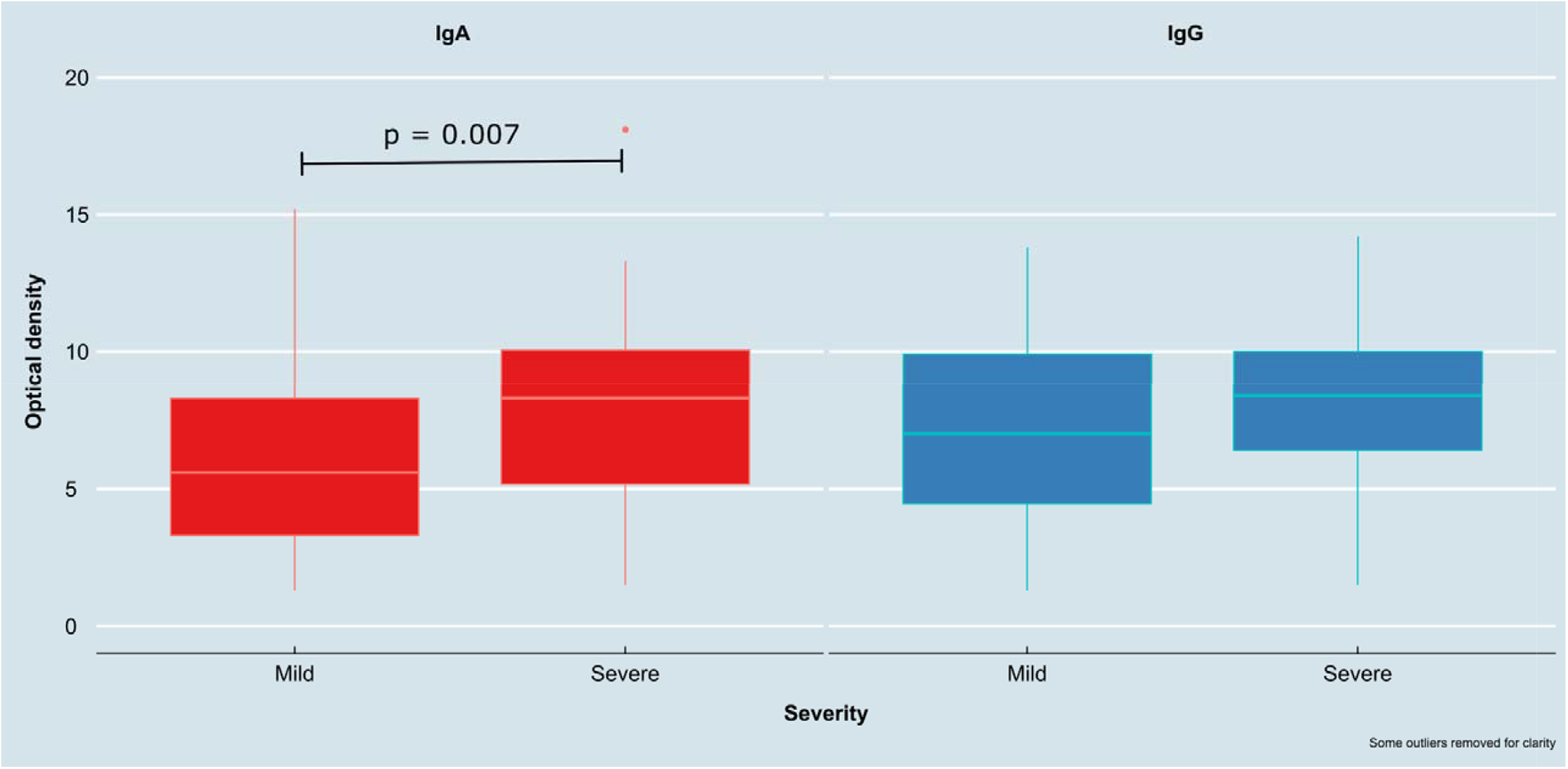
Boxplot showing difference in optical density between severe and mild patients both for IgA and IgG.

### Prospective analysis

Fifty-five patients of our sample were prospectively enrolled; for those patients, 161 blood samples were collected, with an average of 2.93 samples per patient. The average time between the onset of symptoms and the Ig test was 27.3 days. The longest interval between symptoms onset and Ig test was 88 days. Overall, of the 161 samples, 117 tested positive for IgA (72.7%, 95% CI 65.8 – 79.6%), while 107 tested positive for IgG (66.5%, 95% CI 59.2 – 73.8%).

Of those 55 patients, 19 (34.5%) developed a severe form of the disease, while 36 (65.5%) developed a mild form of the disease.

The trend of antibody response showed that both IgA and IgG became detectable between days 5 and 7 from the symptoms onset, peaked between days 21 and 27 and remained stable thereafter, with both IgA and IgG showing a similar trend before day 50. There seemed to be a declining trend for IgA past day 50, while IgG maintained high levels (Figure 4a).

**Figure 4.**
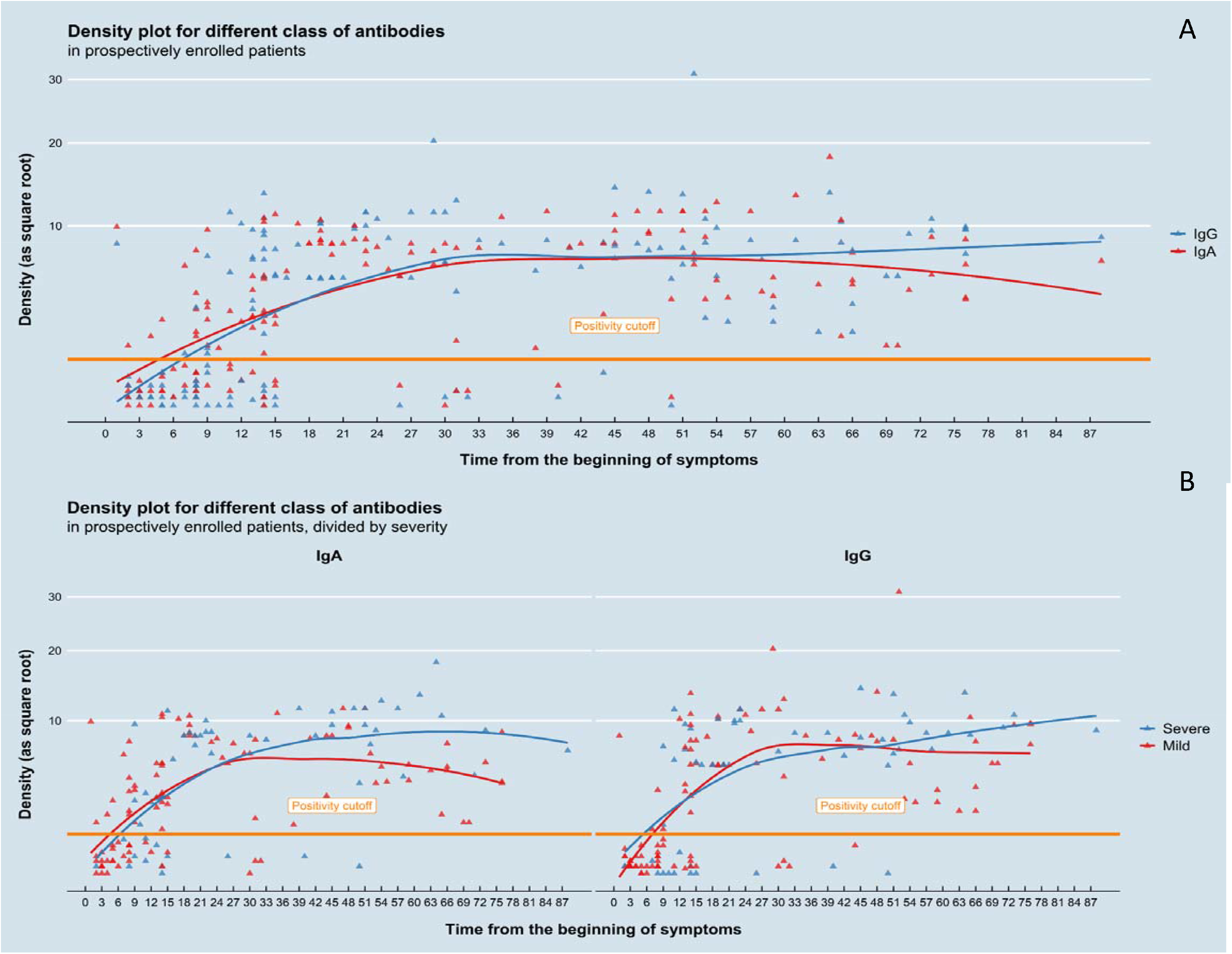
A. Density versus time plot for different Ig class for prospectively enrolled patients. B. Density versus time plot for different Ig class and for different severity of disease in prospectively enrolled patients

After dividing patients according to the severity of disease, peak antibody response appeared to be similar among the two groups. However, there was a more vigorous antibody response both for IgG and IgA in severe patients. Moreover, while IgG tended to stabilize in mild cases, severe cases showed a trend toward more sustained antibody response in time. IgA levels peaked at about the same time in mild and severe patients; however, in mild patients there was a relevant decline after the first month; such a trend was not observable in severe cases (Figure 4b).

## Discussion

Antibody response plays a vital role in viral infections, both for the clearing of the pathogen and for lasting immunity. Serological assays are widely used for the diagnosis of viral infections. However, there is still incomplete knowledge about the kinetic of antibody response to SARS-CoV-2.

Preliminary data reported by Wu et al. out of 175 patients with mild form of disease who did not require hospitalization in ICU highlighted the presence of neutralizing anti-SARS-CoV-2 antibodies 10-15 days after the beginning of the disease. In this paper, however, the authors highlight a rather heterogeneous humoral response in the study population, identifying approximately 30% of patients analyzed with a low antibody titer, therefore suggesting that other immune responses may play an important role. It is not yet known whether these patients are at greater risk of reinfection[29].

Moreover, Zhao et al.[18] in their study conducted among 173 confirmed patients, found out that seroconversion rate was 93,1%, 82,7% and 64,7% for total antibodies (Ab), IgM and IgG respectively, and that median time to Ab, IgM and IgG seroconversion was 11, 12 and 14 days, separately. In this study the seroconversion of Ab was significantly quicker than that of IgM (p = 0.012) and IgG (p<0.001). According to the authors, this is maybe because all isotypes of viral specific antibodies, including not only IgM and IgG but also IgA, can be detected by assay for Ab test.

This evidence supports the hypothesis that other responses to SARS-CoV-2 contribute to virus clearance, and that, to date, we still do not fully know all the mechanisms involved in the immune response to SARS-CoV-2, such as, for example, the role of IgA antibodies against SARS-CoV-2. A similar observation has also been made by Di Giambenedetto et al[19].

Indeed, since the first identification of the SARS-CoV-2, several studies have been published about the kinetic of IgG and IgM response, while little is still known about the IgA response. It is well known that mucosal immunity plays a key role in limiting infections by respiratory pathogens[21], and preliminary data hint that IgA may be an essential part of immune response against SARS-CoV-2 as highlighted from other authors[30–32]. Thus a better understanding of IgA response is needed for SARS-CoV-2.

In our study, IgA response partially resembles IgG response. However, IgA seroconversion seem to happen slightly earlier than IgG. Most patients developed IgA between day 11 and 15, with only small increases of positivity rates thereafter. Conversely, IgG seroconversion seems to occur in a longer time frame, with patients developing antibodies between day 11 and 20. Both for IgA and IgG, the positivity rate remained stable over 80% after day 21 and reached 100% for samples drawn beyond day 60. A recent large-scale seroprevalence study conducted by Gudbjartsson and colleagues showed a similar result, with more than 90% of samples testing positive after the second month since molecular diagnosis of SARS-CoV-2 infection and up to four months[33].

Our data show that there seems to be a difference in the degree of antibody response between mild and severe patients. Specifically, when analyzing all the samples collected, regardless of the time point, there seemed to be a more vigorous antibody response in severe cases compared to mild cases. This difference was more evident for IgA than for IgG. However, we failed to find any difference when blood samples were stratified for time since the onset of symptoms. The same evidence has been highlighted in the studies conducted by Huang and collegues and Hasan Ali and collegues where a higher IgA response was observed in critical and more sever patients[34,35].

Analysis of the prospectively enrolled patients confirmed the findings of the cross-sectional part of the study, with both IgA and IgG showing a similar trend and with IgA appearing slightly earlier than IgG. Moreover, IgA showed a slight decline after the first month. Interestingly, when stratifying for the severity of disease, this decline was evident in mild cases, while no such a decline was observed in severe cases. Unlike IgA, IgG seemed to remain stable in time. Additionally, in mild cases, both the IgA and IgG response seemed to be slightly less vigorous than in severe cases. Also the study conducted by Gudbjartsson and colleagues showed a decline in IgA concentration after the first month[33].

As observed in other infectious diseases, humoral response may be associated with an exaggerated immune response, which, in turn, may be associated with disease progression and with worse prognosis. This mechanism has been also hypothesized for COVID19[36–39]. Indeed, we observed an association between humoral response and severity of disease and such a finding has been reported also by other groups[40]. Interestingly, also the above said-large scale study detected an association between disease severity and strength of humoral response[33].

Based on these premises, it is also possible to speculate that either a higher initial viral load may both be associated with a more vigorous antibody response and a more severe disease or that the lack of viral clearance may be responsible for both sustained inflammation and sustained antibody response (and thus progression of disease to a more severe form).

### Limitations

Our study has several limitations and our results should be interpreted with caution. First, its stated goal is exploratory in nature. Thus, the result of our study should be used only for hypothesis generation, and not to be intended as confirmatory. Where statistical significance has been formally calculated, p values below 0,05 should be interpreted as a signal of a possible association, not as a confirmatory result.

Our study included only hospitalized patients or patients in supervised health care setting who were unable to maintain home isolation, thus generalization to other patients’ populations, especially to asymptomatic patients, should be done with caution. This is particularly true in the case of association between antibody response and severity of the disease. While there seemed to be an association between intensity of IgA response and the severity of the disease when the overall sample was evaluated, we could not find such an association at the single evaluated time frames. This could be due to lack of association or to the fact that the sample size for each time frame was too small to detect such an association. Furthermore, patients who were enrolled in this study were either hospitalized for active disease or had positive swabs for SARS-CoV-2 infection. Thus, they had active antigen stimulation. Our study does not allow us to draw conclusions on patients who have recovered from their disease or have achieved complete viral clearance.

Moreover, in the prospective part of the study, we only collected blood samples while the patients were hospitalized. We were not able to collect blood samples at the prespecified time points after patient’s discharge.

Finally, our institution is a large university hospital that is serving as referral center for the COVID-19 epidemics and our study has been conducted during the initial peak of incidence of COVID-19 in Italy; thus, generalization of our results to other settings or to other time period should be made with caution.

## Conclusion

In summary, IgA and IgG antibody response seem to be closely related. Seroconversion for IgA seems to occur between days 5 and 15 in almost all symptomatic patients. IgG antibody response seems to occur slightly later, peaking at day 20 after symptoms onset. Both IgA and IgG are maintained beyond two months since the beginning of symptoms. There seems to be no correlation between time of antibody response and severity of the disease while such an association is possible for intensity of antibody response. It is possible that a more intense IgA and IgG response is developed in patients with more severe form of the disease. IgA levels seem to decline after one month since the onset of symptoms.

## Data Availability

At the present, raw data are not publicly available

## Declarations

The authors have no funding or conflicts of interest to disclose.

The study was approved by the Ethical Committee of our Institution (ID 3256). Informed consent was obtained from each patient.

## Appendix: Tables and figures

